# Projecting COVID-19 disease severity in cancer patients using purposefully-designed machine learning

**DOI:** 10.1101/2020.08.23.20179838

**Authors:** Saket Navlakha, Sejal Morjaria, Rocio Perez-Johnston, Allen Zhang, Ying Taur

## Abstract

**Background:** Accurately predicting outcomes for cancer patients with COVID-19 has been clinically challenging. Numerous clinical variables have been retrospectively associated with disease severity, but the predictive value of these variables, and how multiple variables interact to increase risk, remains unclear.

**Methods:** We used machine learning algorithms to predict COVID-19 severity in 354 cancer patients at Memorial Sloan Kettering Cancer Center in New York City. Using clinical variables only collected on or before a patient’s COVID-19 positive date (time zero), we sought to classify patients into one of three possible future outcomes: Severe-early (the patient required high levels of oxygen support within 3 days of being tested positive for COVID-19), Severe-late (the patient required high levels of oxygen after 3 days), and Non-severe (the patient never required oxygen support).

**Results:** Our algorithm classified patients into these classes with an AUROC ranging from 70-85%, significantly outperforming prior methods and univariate analyses. Critically, classification accuracy is highest when using a potpourri of clinical variables — including patient demographics, pre-existing diagnoses, laboratory and radiological work, and underlying cancer type — suggesting that COVID-19 in cancer patients comes with numerous, combinatorial risk factors.

**Conclusions:** Overall, we provide a computational tool that can identify high-risk patients early in their disease progression, which could aid in clinical decision-making and selecting treatment options.

## Introduction

At the time of this writing, SARS-CoV-2 infection (COVID-19) continues to exact a substantial toll across a wide range of individuals. Although previous studies have uncovered factors that increase risk of severe COVID-19 infection -- e.g., older age, obesity, or pre-existing heart or lung disease [1–4] -- the clinical course and outcome of patients with COVID-19 illness remains variable and difficult for clinicians to predict. In cancer patients, projecting outcomes can be more complex due to uncertainty regarding cancer-specific risk factors, as well as balancing the risk of an untreated malignancy with the risk of severe infection due to specific anti-neoplastic therapies.

To help clinicians predict COVID-19 severity [5,6], we turned to robust machine learning methods to identify high-risk cancer patients based on their pre-existing conditions and initial clinical manifestations. Prior work using machine learning [7,8] or other analytic techniques has focused on non-cancer patients primarily from China or Italy [9–15]. In this study, we developed a model to predict clinical outcomes (level of oxygen support needed) in cancer patients, using only clinical variables that were available on or before COVID-19 diagnosis (called time zero). Importantly, these variables were selected purposefully, combining both data-driven approaches and expert clinical opinion, and were designed to minimize over-fitting of the model and to increase clinical credibility. We gauged the prospective of this approach to help providers accurately identify cancer patients at greatest risk for impending severe COVID-19 illness, in the hopes of improving outcomes through timely and appropriate interventions.

## Methods

### Study population and clinical variables collected

We analyzed patients admitted to Memorial Sloan Kettering Cancer Center with laboratory-confirmed SARS-CoV-2 (COVID-19) infection from March 10, 2020 (when testing first became available at our institution) to May 1, 2020. The Memorial Sloan Kettering Cancer Center Institutional Review Board granted a Health Insurance Portability and Accountability Act (HIPAA) waiver of authorization to conduct this study.

We aimed to study patients specifically hospitalized for COVID-19 illness by including all patients admitted between 5 days prior, to 14 days after, diagnosis of SARS-CoV-2 infection. COVID-19 patients who were not hospitalized, or who were admitted outside of this window were not included.

An overview of our analysis is shown in Figure 1. For each patient, we extracted and curated 267 clinical variables (Table S1). These included 26 cancer-related variables (e.g., the underlying cancer type, cancer-related medications); 6 demographics variables (e.g., age, sex, race, BMI); 195 variables for preexisting diagnoses (using ICD-9-CM and ICD-10-CM diagnostic code groups; e.g., I1: hypertensive diseases, J4: chronic lower respiratory diseases); 27 clinical laboratory variables (e.g., D-dimer, albumin, lactate dehydrogenase); and 13 radiology variables (e.g., patchy opacities, pleural effusions).

**Figure 1:**
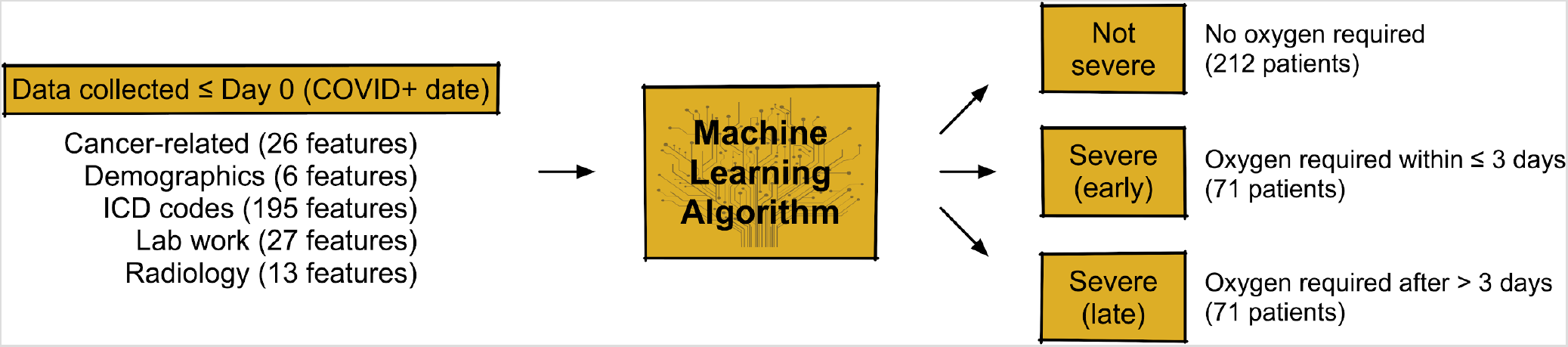
Overview of the study. A) Data for 354 inpatients at Memorial Sloan Kettering Cancer Center were analyzed. For each patient, up to 267 clinical variables were collected, describing the patient’s cancer history, demographics, ICD medical history, laboratory work, and radiology work. Variables were only collected up to the patient’s COVID-19+ date (time zero). B) Variables are inputted into a machine learning algorithm (a random forest classifier), which learns to predict patient outcomes based on interactions between multiple variables. C) Three possible patient outcomes. Of the 354 patients, 212 did not require high levels of oxygen support, 71 required oxygen support within 3 days of being tested positive for COVID-19, and 71 patients required oxygen support after 3 days.

Importantly, we only used clinical variables that were collected on or before a patient’s COVID-19 diagnosis date. For clinical laboratory values, only the most recent value was used. To reduce redundancy, groups of highly correlated variables (Pearson r > 0.90) were removed, and one random variable from the group was kept. Variables could be either mutually exclusive (e.g., indicator variables for a patient having an abnormal vs. a normal X-ray), or overlapping (e.g., having a hematologic cancer vs. leukemia). Overlapping (hierarchical) variables were included to provide the algorithm with multiple resolutions to find discriminating risk factors.

### Defining patient outcomes

Patients were grouped into three possible outcomes based on whether and when they required high levels of oxygenation support, which we defined as oxygen delivered via a nonrebreather mask, high flow nasal cannula (HFNC), bilevel positive airway pressure (BiPAP), or mechanical ventilator. Patients requiring high oxygen support within three days (0 to 3 days relative to COVID-19) were deemed “severe-early”. Patients requiring high oxygen support after three days (4 days after COVID-19 or later) were deemed “severe-late”. Patients not requiring high oxygen (i.e., patients who remained on room air and/or standard nasal cannula) for at least 30 days after COVID-19 were deemed “non-severe”.

Overall, our dataset included 354 inpatients: 212 Non-severe, 71 Severe-early, and 71 Severe-late (Table 1).

**Table 1:** Clinical characteristics (n = 354) and performance statistics. Age and lab values are shown as mean ± std.

|  | Clinical variable | Severe-late | Severe-early | Non-severe |
| --- | --- | --- | --- | --- |
| **Demographics** | # of patients | 71 | 71 | 212 |
|  | Male (%) | 49.3 | 49.3 | 50.0 |
| | Age (years) | 61.9 ± 13.9 | 69.2 ± 12.8 | 59.9 ± 16.8 |
| **Labs** | Absolute Lymphocyte Counts (K/mcL) | 3.57 ± 17.04 | 1.30 ± 2.02 | 2.40 ± 13.78 |
| | D-Dimer (mcg/mL) | 6.58 ± 7.26 | 3.52 ± 2.90 | 2.17 ± 1.72 |
| | Interleukin-6 (pg/mL) | 469.60 ± 816.95 | 116.50 ± 90.25 | 62.62 ± 47.35 |
| | Ferritin (ng/mL) | 1016.36 ± 1045.78 | 983.09 ± 1279.56 | 457.71 ± 537.44 |
| | Platelets (K/mcL) | 171.31 ± 114.04 | 196.04 ± 112.71 | 221.19 ± 124.24 |
| **Cancer-related** | Lymphoma (%) | 11.3 | 5.6 | 1.9 |
|  | Lung (%) | 5.6 | 21.1 | 7.1 |
|  | Leukemia (%) | 19.7 | 8.5 | 5.2 |
| **Diagnosis (ICD)** | I1 - Hypertensive diseases (%) | 52.1 | 71.8 | 56.1 |
|  | I4 - Cardiac disorders (%) | 31.0 | 35.2 | 26.4 |
|  | J4 - Chronic lower respiratory disease (%) | 19.7 | 28.2 | 17.5 |
| **Radiology** | Retic. Opacities (%) | 16.9 | 29.6 | 14.6 |
|  | Effusions (%) | 12.7 | 15.5 | 7.5 |
|  | Airspace Opacity (%) | 32.4 | 81.7 | 37.3 |
| **Performance** | **AUROC (Our method)** | **0.716** | **0.828** | **0.715** |
|  | AUROC (Yan et al.) | 0.461 | 0.638 | 0.511 |
|  | AUROC (Huang et al.) | 0.596 | 0.628 | 0.596 |
|  | **Avg. Precision (Our method)** | **0.365** | **0.576** | **0.780** |
|  | Avg. Precision (Yan et al.) | 0.194 | 0.316 | 0.607 |
|  | Avg. Precision (Huang et al.) | 0.286 | 0.317 | 0.682 |

### Machine learning algorithms and validation

To predict patient outcomes, we employed a random forest ensemble machine learning algorithm, consisting of multiple independent classifiers, each trained on different subsets of training variables [16]. These classifiers collectively estimate the patient’s most likely outcome. Our random forest model consisted of 500 decision trees, trained using the information gain criterion, and each with a maximum depth of 10 decision nodes and a minimum of 1 sample per leaf.

The model was evaluated using 10-fold stratified cross-validation, in which 90% of the dataset (approximately, 318 patients) were used to train the model, and the remaining 10% of the dataset (36 patients) were used to test the model. This process was repeated 10 times, such that each subject was assigned to the test set exactly once. This procedure also ensured that each fold had a class (outcome) distribution that approximately matched that of the complete dataset. We report area under the receiver operating characteristic (AUROC) and average precision scores for each class separately using a one-vs.-rest classification scheme [17].

The importance of each clinical variable towards performance was assessed using permutation testing [16], in which values for each variable (column) were randomly permuted over the observations and then model performance was re-assessed using cross-validation; the drop in performance was used as a measure of the variable’s importance.

### Comparison of performance to prior work

Previous machine learning studies have reported impressive performance predicting COVID-19 outcomes for non-cancer patients using only a few clinical variables. For example, Yan et al. [7] (*Nature Mach. Intell*., 2020) report 90+% performance using just three variables (lactate dehydrogenase, C-reactive protein, and absolute lymphocyte count). Huang et al. (*Lancet*, 2020) reported statistical significance for 10 clinical variables (white blood cell count, absolute neutrophil count, absolute lymphocyte count, prothrombin time, D-dimer, albumin, total bilirubin, lactate dehydrogenase, troponin I, and procalcitonin). Other studies also used many of the same clinical variables [10,13,14]. For a fair comparison, and to test whether variables previously identified as important could also well-predict outcomes for cancer patients, we trained two random forest classifiers on our dataset using only the variables used by Yan et al. and Huang et al., respectively.

### Experimental setup and rationale

Wynants et al. [8] recently reviewed 16 prognostic models for predicting COVID-19 severity and concluded that every study had a high or unclear risk of bias. To try and minimize bias in our analytic approach, we followed three guidelines suggested by the authors:

a. *Practices to reduce model over-fitting*. We used cross-validation, a standard practice in machine learning, to test how well a trained model can predict outcomes on patients it has never seen before. Evaluating models in this way helps to ensure that predictive patterns learned by the model can generalize to new patients whose outcomes are unknown.
b. *Using a hybrid of expert clinical opinion and data-driven approaches to select variables*. The authors of our study include both clinicians and computer scientists, who collaborated closely to home-in on a set of relevant clinical variables. As an example, using a completely data-driven approach, we found that a class of medications, atypical antipsychotics, correlated highly with disease severity; in fact, including these medications in our model would have increased our reported results by ∼4-5%. However, these medications are frequently given to elderly patients with dementia, and we felt these medications were very unlikely to directly cause severe COVID-19, and far more likely to be confounded by age and/or functional status. So, we removed this variable and instead decided “age” was the more appropriate and general risk factor. Thus, we began with a purely data-driven approach to identify candidate variables, and then iteratively eliminated those that seemed tenuous from a clinical perspective. Our final model was trained using only 55 of the 267 variables (Table S1).
c. *Only including patients who had sufficient time to experience their outcome by the end of the study*. We evaluated hospitalized patients diagnosed with COVID-19 from March 10 to May 1, 2020, and evaluated outcomes from March 10 until May 15, 2020, to ensure at least two weeks of follow-up for all patients.

## Results

From March 10, 2020 to May 1, 2020, there were 354 inpatients at Memorial Sloan Kettering Cancer Center in New York City, the heart of the COVID-19 pandemic. Below, we test several models for predicting disease severity in this cancer patient cohort.

### Univariates and bivariates weakly correlate with COVID-19 patient outcomes

Figure 2A-F shows that neither of six clinical variables commonly associated with COVID-19 severity (age, C-reactive protein, D-dimer, albumin, lactate dehydrogenase, BMI) are by themselves able to discriminate among the three patient outcomes. Some laboratory variables can only stratify between non-severe and severe-early patients (e.g., Figure 2B, C-reactive protein), indicating that these labs may only be valuable for prognosing immediate risk as opposed to future risk. Others laboratory variables may be more discriminative but were only available for a fraction of patients at time zero (e.g., Figure 2C, D-dimer). Overall, none of the variables we tested were significantly different between all three outcome groups (non-severe, severe-early, severe-late).

**Figure 2:**
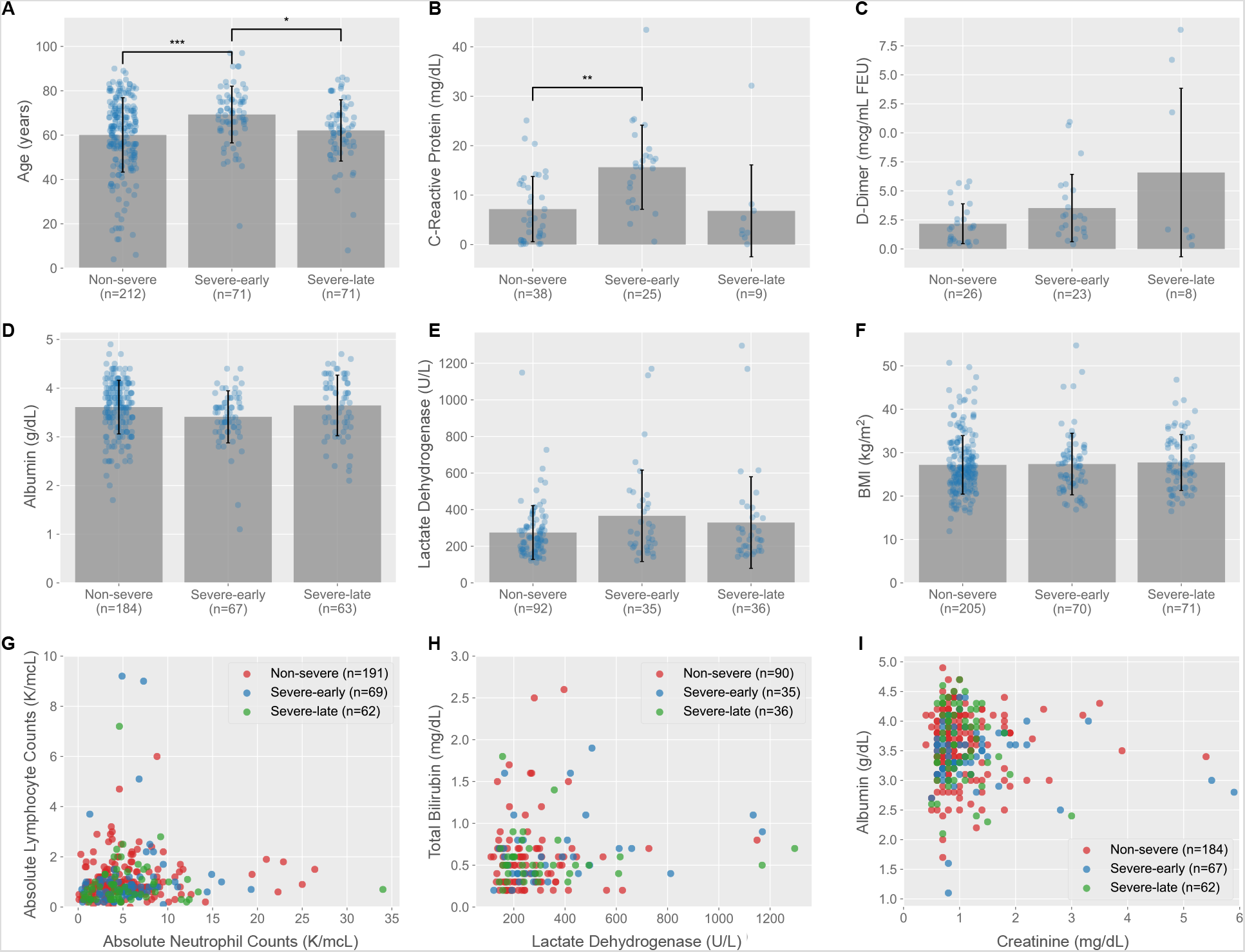
Individual clinical variables weakly correlate with patient outcomes. A-F) Each panel shows a variable (y-axis) grouped by patients in each of the three outcomes (x-axis). The number of patients (n) for which the variable was measured is shown for each group. For example, there were 212 non-severe patients and their average age was 59.9 years old. Each bar shows average; error bars show standard deviation. A) Age, B) C-reactive protein, C) D-dimer, D) Albumin, E) Lactate dehydrogenase, F) BMI. G-I) Each panel shows an interaction between two variables (x and y axes). Each patient is represented by a colored dot (red = non-severe, blue = severe-early, green = severe-late). * = P < 0.01, ** = P < 0.001, *** = P < 0.0001, Welch’s two-sample T-test.

We next looked at whether interactions between two variables could be used to increase prediction accuracy. While there are hundreds of pairs of variables to test, Figure 2G-I shows three representative plots using pairs of commonly used labs, none of which show any visually clear clustering of patients by outcome (i.e., clustering of the same colored dots together).

### Improved prediction using machine learning

To test if a combinatorial approach, which takes interactions between numerous risk factors into account, may improve projections of COVID-19 severity, we trained an ensemble machine learning algorithm using a wide range of clinical variables (as described in Methods). Clinical variables included those related to the patient’s underlying cancer diagnosis and treatment, laboratory work, radiological work, pre-existing diagnoses (ICD code history), and demographics. We validated our model using cross-validation, in which a portion of the patients were used to train the model, and the model was then evaluated on the remaining or left-out patients, whose outcomes are known but are never provided to the model.

Our model accurately predicted outcomes for COVID-19 cancer patients who required high levels of oxygen support within 3 days of COVID-19 diagnosis (AUC = 0.828 for severe-early patients; Figure 3A). The model achieved fair accuracy in the more challenging instances of predicting severity that occurs after 3 days (AUC = 0.716 for severe-late patients) or that never occurs during the length of the patient’s disease (AUC = 0.715 for non-severe patients). The model maintains an AUC of greater than 0.8 if “severe-early” was defined as all patients that required oxygen support within 4 days of diagnosis (instead of 3 days), but performance then begins to drop at longer time horizons: AUC = 0.821 for ≤ 4 days (81 patients); AUC = 0.791 for ≤ 5 days (88 patients); and AUC = 0.727 for ≤ 6 days (99 patients). These results suggest that prediction is only reliable within a 3-4 day window from the time of diagnosis.

**Figure 3:**
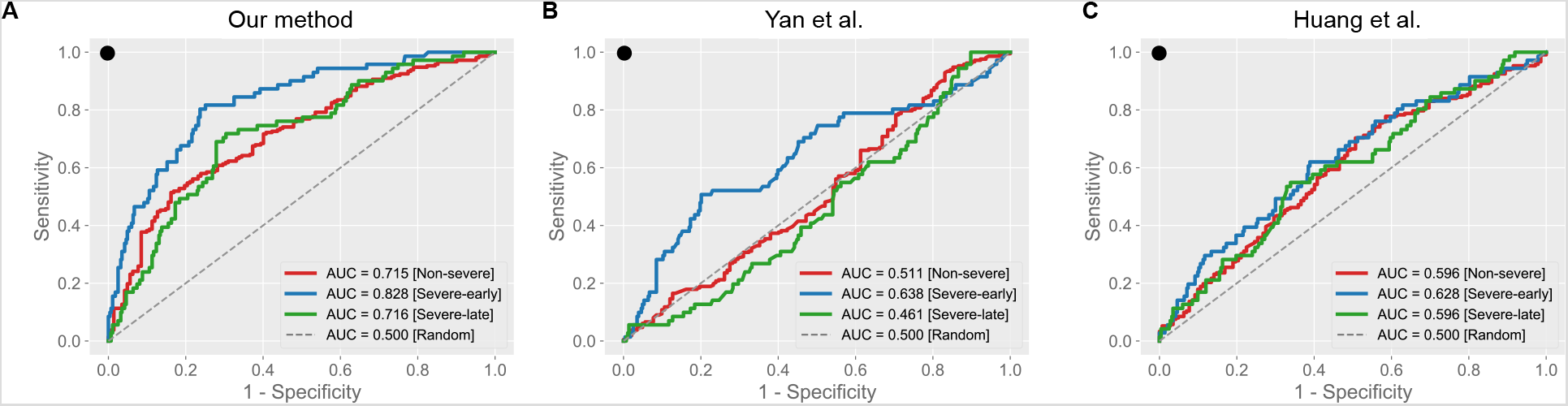
Machine learning algorithms improve COVID-19 outcome prediction in cancer patients. AUROC plots for A) Our method, B) Yan et al. (2020), and C) Huang et al. (2020). AUROCs are reported for each class separately using a one-vs.-rest evaluation scheme. Diagonal dotted line shows random prediction (AUROC of 0.500). Perfect prediction lies at the upper left of the plot (black dot).

Prior work has reported that a small set of clinical variables can serve as a robust “signature” of COVID-19 disease severity [1,7] (Methods). However, we found significantly worse performance using these variables (Figure 3B-C). For example, for severe-early patients, Yan et al. (3 variables) and Huang et al. (10 variables) achieved AUCs of 0.638 and 0.628, compared to 0.828 for our method. Similarly, for non-severe patients, the two studies achieved AUCs of 0.511 and 0.596, compared to 0.715 for our method. We also computed average precision scores (a summary statistic of the precision-recall curve) and found similar gains for our method compared to prior works (Table 1).

### Identifying variable and multi-variable interactions that are useful for predicting patient outcomes

Figure 4A shows the top 30 variables that were most discriminative in classifying patient outcomes. These were variables which, if effectively removed from the analysis, would result in a drop in performance (Methods). For example, platelets, ferritin, and AST (aspartate aminotransferase) were the three most important individual labs. Because we used dozens of variables, and many variable combinations may be correlated, we do not expect the loss of one or a few variables to make a significant difference in performance. Nonetheless, many of these variables have been previously identified in the COVID-19 literature (e.g., interleukin 6 [18,19], C-reactive protein [20], D-dimer). Interestingly, there are also variables the model used that are less discussed in the literature, including ferritin [12,21]. Our study also highlights the importance of variables related to cancer diagnoses and treatments on COVID-19 severity; for example, whether the patient had leukemia or lung cancer was particularly discriminative.

**Figure 4:**
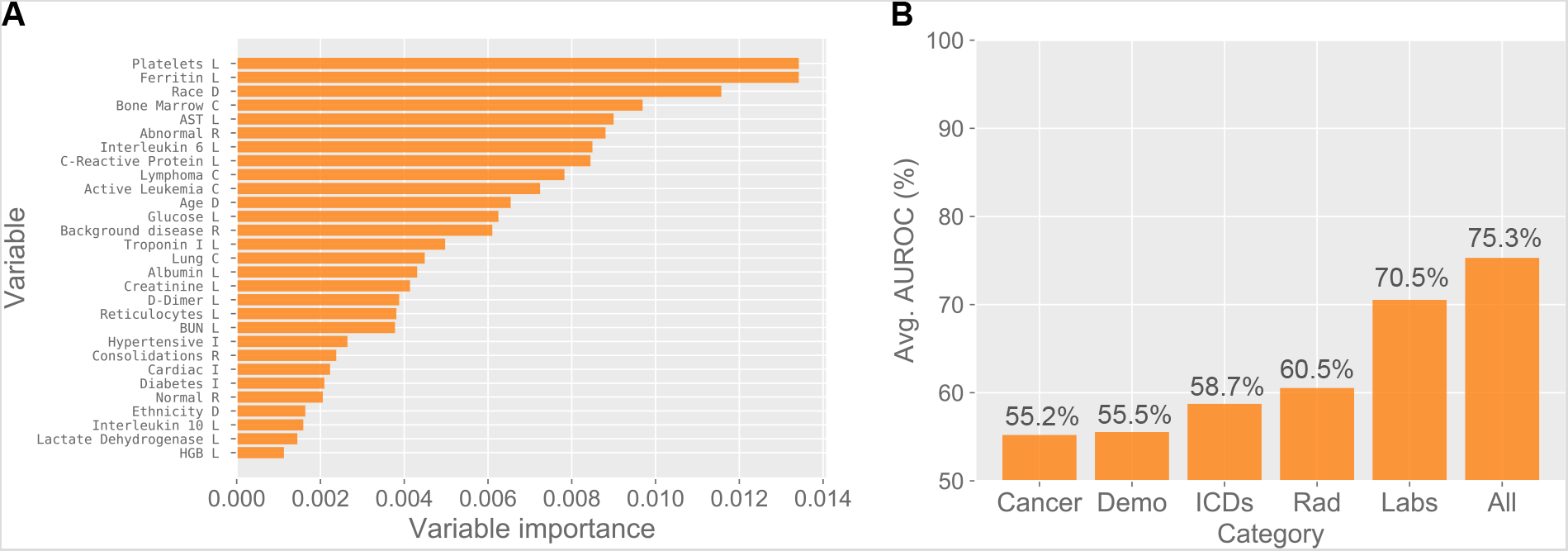
Important clinical variables identified by the model. A) The top 30 variables (y-axis) and their importance (x-axis), defined using permutation testing. The category of each variable is listed next to its name: C = Cancer-related, D = Demographics, I = ICD codes, R = Radiology, L = Laboratory. B) The performance of the classifier (y-axis) when trained using variable from each category separately. For example, using only radiology variables, the random forest classifier achieved an AUROC, averaged over all three classes, of 0.605. “All” shows the combination of all variables, achieving an average AUROC of 0.753.

Variables from all five categories (cancer-related, demographics, ICD codes, laboratory work, radiological work) are represented in Figure 4A, highlighting how each clinical category contributes complementary information towards projecting COVID-19 severity. Indeed, classifying patient outcomes using variables from each category individually reduces accuracy compared to when using all variables together (Figure 4B). For example, training the model using only cancer variables produced an average AUROC of only 55.3%. On the other hand, using all variables except cancer-related variables dropped performance by 6.9%. The former means that the underlying cancer type, by itself, is not a very valuable predictor, but the latter suggests that when the cancer type is combined with clinical variables from other categories, its contribution becomes more pronounced and is unique. Similarly, using only radiology variables produced an average AUROC of 60.5%, and using all variables except for radiology variables dropped performance by 7.6%.

## Discussion

We used machine learning algorithms to identify clinical variables predictive of severe COVID-19 illness in cancer patients at time zero. We achieved an AUC ranging from 70-85%, with high performance for classifying patients with an immediate risk of decompensation (severe-early, ≤ 3 days), and fair performance for patients with less immediate risks (severe-late, > 3 days) or no risk at all (not-severe). Our tool is designed to complement (not replace) a clinician’s experience and judgement and may be most helpful to untangle complex interactions among multiple risk factors.

Following the guidelines of Wynant et al. [8], we combined data-driven variable selection with expert clinical opinion to reduce overfitting and minimize bias in the model. Had we included all variables, our model’s performance would increase by at least 5%, but we deliberately did not report these results and instead opted to build a model with more clinical credibility. In addition, our study was meant to tackle two real-world challenges in treating COVID-19 patients. First, we used the time of COVID-19 diagnosis (time zero) as a landmark, and we only provided to our model data available on or before time zero in order to represent the information available to providers at the time of presentation and diagnosis. As a result, there may be a lack of consistency in what clinical variables are available for the model to use. For example, even though D-dimer are commonly associated with COVID-19 severity [22], very few of our patients (16.1%, 56/354) had available D-dimer labs on the date of their COVID-19 diagnosis. Second, patients enter the hospital at different points in their disease progression, and we did not attempt to correct for these differences. A useful model, we reasoned, needs to deal with this lack of synchronicity to be practical.

There are several advantages and disadvantages to the machine learning approach taken here. On the plus side, automated models can help evaluate a large pool of clinical variables as risk factors for disease severity, with potential to go beyond conventional modelling approaches, which are generally limited to evaluation of only a handful of variables. Further, evaluating the model using cross-validation reduces the probability of overfitting and highlights a model’s prognostic ability. On the downside, the model seeks variables that are correlated with patient outcomes, and these variables are not necessarily causal drivers of the disease. For example, corticosteroids given to severe COVID-19 patients are known to affect blood glucose levels, and our model makes no attempt to distinguish the directionality of the interaction between the two. We attempted to overcome this by using a hybrid of expert clinical opinion and data-driven approaches to select variables in a purposeful manner, though it remains a challenge to know how to weigh the importance of clinical experience and data.

Moving forward, several challenges remain in bringing clinical machine learning to the bedside for COVID-19 treatment. First, we analyzed a modestly-sized dataset of 354 cancer patients; larger, more comprehensive datasets of cancer patients are needed to test the true generality of our approach. Second, better algorithms are needed to forecast future outcomes (severe-late and non-severe); e.g., time-series analyses of how clinical variables change over time may provide one avenue forward. Third, models should aid clinicians in the real-time process of deciding which diagnostic tests to order on a patient based on the putative discriminative power of the test results. Ideally, models would interact with clinicians in a back-and-forth manner to home-in on the clinical variables most critical for accurate forecasting [23].

To better prepare us for the next outbreak –- be it a second wave of COVID-19 or something else altogether -- we hope that physicians, epidemiologists, and computer scientists will continue working together to understand and build useful models to predict an individual’s susceptibility to disease.

## Data Availability

Data will be made available following publication.

## Acknowledgements

The authors thank Anthony Daniyan and Sham Mailankody for help with chart-reading of cancer variables. SN thanks funding from the Simons Center for Quantitative Biology at Cold Spring Harbor Laboratory.

---

**Table S1**: List of all 267 variables. Bolded features are the subset of 55 used in our analysis.

1. Demographics (6): Basic patient information.

Sex, **Race, Ethnicity, Age, Smoker, BMI**
2. Labs (27): Laboratory work variables.

**ALT**, APTT, **AST**, Absolute Lymphocytes, **Absolute Neutrophils, Absolute Reticulocytes, Albumin, BUN**, Total Bilirubin, **C-Reactive Protein, Creatinine, D-Dimer, Erythrocyte Sedimentation Rate, Ferritin, Glucose, HGB, Interleukin 1 beta, Interleukin 10, Interleukin 6, Lactate Dehydrogenase, PT, Platelets, Procalcitonin, Troponin I**, WBC, E6, **ANC-ALC-ratio**
3. Cancer-related (26): Each variable is binary, indicating whether the patient has the cancer or is taking a cancer-related medication.

**Bladder, Bone Marrow, Breast**, Conn, Endometrium, **GI, GU**, Kidney, **Leukemia, Lung, Lymph Nodes, NHL (Non-Hodgkins Lymphoma), PCD (Plasma Cell Dycrasias)**, Pancreas, Prostate, Rectum, Sigmoid Colon, Skin, Thyroid
Active Cancer: Whether the patient received radiation, chemotherapy, or surgical intervention in the last 6 months.
**Check-point inhibitors**: Whether the patient is taking one of: pembrolizumab, nivolumab, atezolizumab, avelumab, durvalumab, ipilimumab.
**Corticosteroids**: Whether the patient is chronically on prednisone, prednisolone, or dexamethasone. Chronic is defined as > 5 mg of prednisone (or equivalent) for 5 days.
Group: **Hematologic, HCT, Solid**
4. ICD codes for pre-existing diagnoses (195): The first two characters of the ICD-10 code. Each variable is binary, indicating whether the ICD code was assigned to the patient. Examples include:

**E1**: diabetes mellitus
**I1**: hypertensive diseases
**I4**: cardiac disorders
**J4**: chronic lower respiratory diseases
**Major Surgery**: Whether the patient had a major surgery in the last 30 days that required general anesthesia.
5. Radiology (13): Patient’s X-rays were classified as “normal”, “abnormal” or “indeterminate”. When available, current studies were compared to prior to ensure findings were new. If patients prior X-rays demonstrated findings, these were classified as having “background disease”. If no findings were detected on the X-ray or no new findings were noted when compared to prior, the study was determined to be “normal”. Studies with extensive background metastasis, large neoplasms, effusions or post treatment changes, which would significantly obscure new findings, were classified as “indeterminate”. A study was determined as “abnormal” if airspace and/or reticulonodular opacities were noted. Airspace opacities were divided into patchy opacities and segmental or lobar consolidations. Findings were further classified as “unilateral” or “bilateral” and as predominantly in the inferior lobes (“gradient”) or diffuse involving upper lobes. In addition, pleural effusions were recorded and classified by amount (small, moderate or large).

**Normal**
**Abnormal**
**Indeterminate**
**Background disease**
**Unilateral**
Bilateral
Airspace Opacities
Patchy Opacities
**Consolidations**
Gradient (1. basilar, 2. upper and/or lower)
**Reticulonodular Opacities**
**Effusions**
Amount (1. small, 2 moderate, 3. large)

## References

1. Huang C, Wang Y, Li X, Ren L, Zhao J, Hu Y, et al. Clinical features of patients infected with 2019 novel coronavirus in Wuhan, China. Lancet. 2020;395: 497–506.

2. Du R-H, Liang L-R, Yang C-Q, Wang W, Cao T-Z, Li M, et al. Predictors of mortality for patients with COVID-19 pneumonia caused by SARS-CoV-2: a prospective cohort study. Eur Respir J. 2020;55. doi:10.1183/13993003.00524-2020

3. Jain V, Yuan J-M. Predictive symptoms and comorbidities for severe COVID-19 and intensive care unit admission: a systematic review and meta-analysis. Int J Public Health. 2020. doi:10.1007/s00038-020-01390-7

4. Li B, Yang J, Zhao F, Zhi L, Wang X, Liu L, et al. Prevalence and impact of cardiovascular metabolic diseases on COVID-19 in China. Clin Res Cardiol. 2020;109: 531–538.

5. Richardson S, Hirsch JS, Narasimhan M, Crawford JM, McGinn T, Davidson KW, et al. Presenting Characteristics, Comorbidities, and Outcomes Among 5700 Patients Hospitalized With COVID-19 in the New York City Area. JAMA. 2020. doi:10.1001/jama.2020.6775

6. Robilotti EV, Babady NE, Mead PA, Rolling T, Perez-Johnston R, Bernardes M, et al. Determinants of COVID-19 disease severity in patients with cancer. Nat Med. 2020. doi:10.1038/s41591-020-0979-0

7. Yan L, Zhang H-T, Goncalves J, Xiao Y, Wang M, Guo Y, et al. An interpretable mortality prediction model for COVID-19 patients. Nat Mach Intell. 2020;2: 283–288.

8. Wynants L, Van Calster B, Collins GS, Riley RD, Heinze G, Schuit E, et al. Prediction models for diagnosis and prognosis of covid-19 infection: systematic review and critical appraisal. BMJ. 2020;369: m1328.

9. Fabio C, Antonella C, Patrizia R-Q, Francesco DC, Annalisa R, Laura G, et al. Early predictors of clinical outcomes of COVID-19 outbreak in Milan, Italy. Clin Immunol. 2020; 108509.

10. Wang D, Hu B, Hu C, Zhu F, Liu X, Zhang J, et al. Clinical Characteristics of 138 Hospitalized Patients With 2019 Novel Coronavirus-Infected Pneumonia in Wuhan, China. JAMA. 2020. doi:10.1001/jama.2020.1585

11. Yang X, Yu Y, Xu J, Shu H, Xia J’an, Liu H, et al. Clinical course and outcomes of critically ill patients with SARS-CoV-2 pneumonia in Wuhan, China: a single-centered, retrospective, observational study. Lancet Respir Med. 2020;8: 475–481.

12. Zhou F, Yu T, Du R, Fan G, Liu Y, Liu Z, et al. Clinical course and risk factors for mortality of adult inpatients with COVID-19 in Wuhan, China: a retrospective cohort study. Lancet. 2020;395: 1054–1062.

13. Ruan Q, Yang K, Wang W, Jiang L, Song J. Clinical predictors of mortality due to COVID-19 based on an analysis of data of 150 patients from Wuhan, China. Intensive Care Med. 2020;46: 846–848.

14. Guan W-J, Ni Z-Y, Hu Y, Liang W-H, Ou C-Q, He J-X, et al. Clinical Characteristics of Coronavirus Disease 2019 in China. N Engl J Med. 2020;382: 1708–1720.

15. Tang N, Li D, Wang X, Sun Z. Abnormal coagulation parameters are associated with poor prognosis in patients with novel coronavirus pneumonia. J Thromb Haemost. 2020;18: 844–847.

16. Breiman L. Random Forests. Machine Learning. 2001;45: 5–32.

17. Bishop CM. Pattern Recognition and Machine Learning. Springer; 2006.

18. Wang Z, Yang B, Li Q, Wen L, Zhang R. Clinical Features of 69 Cases with Coronavirus Disease 2019 in Wuhan, China. Clin Infect Dis. 2020. doi:10.1093/cid/ciaa272

19. Sun D, Li H, Lu X-X, Xiao H, Ren J, Zhang F-R, et al. Clinical features of severe pediatric patients with coronavirus disease 2019 in Wuhan: a single center’s observational study. World J Pediatr. 2020. doi:10.1007/s12519-020-00354-4

20. Wang G, Wu C, Zhang Q, Wu F, Yu B, Lv J, et al. C-Reactive Protein Level May Predict the Risk of COVID-19 Aggravation. Open Forum Infect Dis. 2020;7: ofaa153.

21. Shoenfeld Y. Corona (COVID-19) time musings: Our involvement in COVID-19 pathogenesis, diagnosis, treatment and vaccine planning. Autoimmun Rev. 2020;19: 102538.

22. Lippi G, Favaloro EJ. D-dimer is Associated with Severity of Coronavirus Disease 2019: A Pooled Analysis. Thrombosis and haemostasis. 2020. pp. 876–878.

23. Settles B. Active Learning. Synthesis Lectures on Artificial Intelligence and Machine Learning. 2012. pp. 1–114. doi:10.2200/s00429ed1v01y201207aim018

